# Clinical importance of reporting SARS-CoV-2 viral loads across the different stages of the COVID-19 pandemic

**DOI:** 10.1101/2020.07.10.20149773

**Authors:** M. Moraz, D. Jacot, M. Papadimitriou-Olivgeris, L. Senn, G. Greub, K. Jaton, O. Opota

## Abstract

On April 25^th^, corresponding to the first deconfinement phase after the end of the lockdown in Switzerland, a universal admission screening strategy for COVID-19 was introduced in our hospital. All patients, including asymptomatic patients were tested for SARS-CoV-2 by quantitative reverse transcription polymerase chain reaction (RT-PCR). In addition to a qualitative answer, providing viral load values to the RT-PCR results not only helped the clinician to evaluate the stage of the infection but addressed patient contagiousness and guided infection control decisions. Here, we discuss the importance of reporting viral load values when a shift from a symptomatic to a universal screening strategy was performed.

---

Since the beginning of the COVID-19 pandemic, SARS-CoV-2 quantitative reverse transcription polymerase chain reaction (RT-PCR) was used as a crucial diagnostic tool for patient care, hospital hygiene decisions and health authorities policy (1). In addition to qualitative “yes or no” answer, RT-PCR can provide quantitative values based on cycle threshold values (Cts) and their corresponding calculated viral loads. However, several studies have reported i) that viral loads may vary significantly among patients, clinical specimens and during the infection (Jacot et al., submitted) and ii) that RT-PCRs could remain positive up to five weeks after onset of symptoms (2). Between the beginning of March and May 17^th^, our molecular diagnostic laboratory located in a tertiary care university hospital (Lausanne, Switzerland), performed more than 34’000 SARS-CoV-2 RT-PCRs using three different platforms: our high throughput automated molecular diagnostic platform (MDx platform) (3), the cobas SARS-CoV-2 test and the GeneXpert Xpress SARS-CoV-2. All three platforms showed comparable performance with correlation of the Cts values (Jacot et al., submitted and Opota et al., submitted).

Reporting Ct values can be beneficial for the interpretation of RT-PCR results (4). Such information can be used i) by the laboratory as an internal quality assessment tool, ii) by clinicians to evaluate the progression of the infection (especially in lower respiratory tract specimens) or iii) to address patient contagiousness and hence to guide infection control decisions. Regarding the latter application, the benefit of reporting the Ct value appeared of utmost importance when a universal admission or pre-intervention screening strategy including asymptomatic patients was introduced in our hospital from April 25^th^, as supported by the analysis of 32’005 RT-PCR results obtained in our laboratory. Overall, we observed higher Ct values (lower viral loads) in patients screened after April 25^th^ than for patients, mostly symptomatic, screened during the “epidemic period” (Fig. 1). This abrupt change was confirmed by looking only at a 2 weeks period just prior to the universal screening. The GeneXpert SARS-CoV-2 test was broadly used during this period, detecting the SARS-CoV-2-specific N2 region (encoding for viral nucleoprotein N2) and the E-gene (encoding for a protein of the envelope, Cts for the E-gene were used as they correlated with the other two platforms). Among the very high Cts, we obtained several results positive only for N2 suggesting a very low viral load at the detection limit of the GeneXpert assay (5, 6). By retesting these specimens with other RT-PCR platforms and reviewing clinical data, we could demonstrate that these N2 only positive results corresponded to true detection of viral RNA. This reinsured us on the specificity of the test when the percentage of positive results dramatically decreased due to the reduced virus circulation and to the strategy of universal screening, including asymptomatic patients. Since April 25^th^, we reported the viral load for all the SARS-CoV-2 PCR with a lower threshold of 1’000cp/ml as “positive result <1’000cp/ml” (Ct >37). When only the N2 gene was positive, the reported result was “very low positive with impossible quantification” as viral load quantification was performed on the E gene.

**Figure 1:**
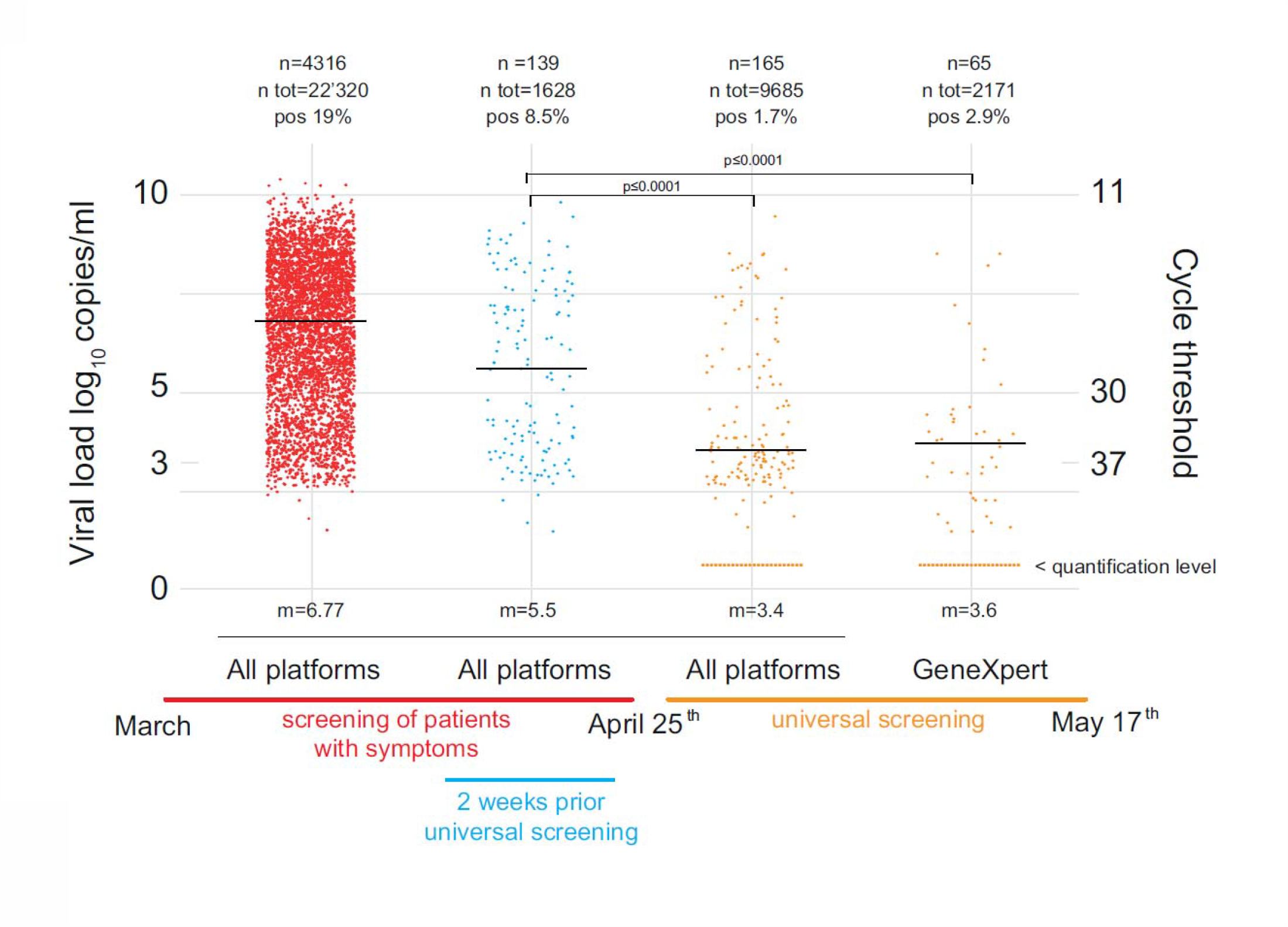
Median viral load value of positive SARS-CoV-2 RT-PCR compared across two periods of the COVID-19 pandemic. Median viral load value of positive SARS-CoV-2 RT-PCR was compared across two periods: the “epidemic period” (left), during which mainly symptomatic patients were screened, and the “post-epidemic period” (right) when all patients were tested on hospital admission. A sharp decrease of median viral load (often <1’000 cp/ml) values was observed during the universal screening period when most subjects tested were asymptomatic. Patient samples analyzed using the GeneXpert test and showing only an N positive PCR are displayed below the quantification limit. Cts were converted to viral loads using the formula −0.27Ct+13.04 generated using purified viral RNA, kindly provided by the Institute of Virology of the University of Berlin, la Charité (8). Significance of viral load decrease was assessed using a parametric paired t-test with p-values ≤ 0.0001. m= Ct value median, n= number of positive samples, n tot= total number of tests, pos= percentage of positive tests.

In addition to clinical data, providing quantitative viral loads values can help epidemiologists for infection control strategy. According to the Centers for Disease Control and Prevention, transmission-based precautions might be reconsidered either when 10 days have passed since symptoms first appeared or following at least two consecutive negative RT-PCR results (7). However, even with the complete resolution of symptoms, some patients can have a prolonged positive test result (2). We observed patients’ specimens positive results although with very high Cts, up to 5 weeks after infection. This might correspond to nonviable virus and/or to patients with very low transmission potential. While many pre-analytical issues can affect the result of any respiratory virus RT-PCRs, our data also highlight the importance of viral load quantification for the interpretation of SARS-CoV-2 RT-PCRs for the care of patients with positive results, especially at the end of viral disease.

## Data Availability

All the data reffered to in the manuscript are available

## Acknowledgements

We would like to deeply thank all the staff of the Institute of Microbiology of the Lausanne University Hospital. In particular we would like to thank all the biomedical technicians of the molecular diagnostic laboratory for their incredible work and support during the pandemic.

## References

1. Deng SQ, Peng HJ. Characteristics of and Public Health Responses to the Coronavirus Disease 2019 Outbreak in China. J Clin Med. 2020;9(2).

2. Xiao AT, Tong YX, Zhang S. Profile of RT-PCR for SARS-CoV-2: a preliminary study from 56 COVID-19 patients. Clin Infect Dis. 2020.

3. Greub G, Sahli R, Brouillet R, Jaton K. Ten years of R&D and full automation in molecular diagnosis. Future Microbiol. 2016;11(3):403–25.

4. Tom MR, Mina MJ. To Interpret the SARS-CoV-2 Test, Consider the Cycle Threshold Value. Clin Infect Dis. 2020.

5. Moran A, Beavis KG, Matushek SM, Ciaglia C, Francois N, Tesic V, et al. The Detection of SARS-CoV-2 using the Cepheid Xpert Xpress SARS-CoV-2 and Roche cobas SARS-CoV-2 Assays. J Clin Microbiol. 2020.

6. Loeffelholz MJ, Alland D, Butler-Wu SM, Pandey U, Perno CF, Nava A, et al. Multicenter Evaluation of the Cepheid Xpert Xpress SARS-CoV-2 Test. J Clin Microbiol. 2020.

7. CDC. Discontinuation of Isolation for Persons with COVID-19 Not in Healthcare Settings. 2020.

8. Corman VM, Landt O, Kaiser M, Molenkamp R, Meijer A, Chu DK, et al. Detection of 2019 novel coronavirus (2019-nCoV) by real-time RT-PCR. Euro Surveill. 2020;25(3).

